# Reconciling the efficacy and effectiveness of masking on epidemic outcomes

**DOI:** 10.1101/2023.05.10.23289803

**Authors:** Wan Yang, Jeffrey Shaman

## Abstract

Mask wearing in public settings has been broadly implemented as a means to mitigate the COVID-19 pandemic. However, the reported effectiveness of masking has been much lower than laboratory measures of efficacy, and this large discrepancy has cast doubt on the utility of masking. Here, we develop an agent-based model that comprehensively accounts for individual masking behaviors and infectious disease dynamics, and test the impact of masking on epidemic outcomes. Using realistic inputs of mask efficacy and contact data at the individual level, the model reproduces the lower effectiveness as reported in randomized controlled trials. Model results demonstrate that transmission within households, where masks are rarely used, can substantially lower effectiveness, and reveal the interaction of nonlinear epidemic dynamics, control measures (e.g., masking and social distancing), and potential measurement biases. Overall, model results show that, at the individual level, consistent masking can reduce the risk of first infection, and, over time, reduce the frequency of repeated infection. At the population level, masking can provide direct protection to mask wearers, as well as indirect protection to non-wearers, collectively reducing epidemic intensity. These findings suggest it is prudent for individuals to use masks during an epidemic, and for policy makers to recognize the less-than-ideal effectiveness of masking when devising public health interventions.

**Significance statement:** During the COVID-19 pandemic, mask wearing in public settings has been a key control measure. However, the low effectiveness reported for masking has cast doubt on its validity. Here, we develop an agent-based model to interrogate influencing factors. Using realistic inputs of mask efficacy and contact data, the model reproduces the lower effectiveness reported in real-world settings. Testing shows that transmission within-household where masks are rarely used can substantially lower effectiveness. Nonetheless, the model results support the effectiveness of masking at both the individual and population levels, albeit at less-than-ideal levels. Overall, these findings indicate it is prudent for individuals to use masks during an epidemic, and for policy makers to recognize the less-than-ideal effectiveness of masking when devising interventions.

## INTRODUCTION

During the coronavirus disease 2019 (COVID-19) pandemic, universal mask wearing in public settings has been implemented in many places throughout the world as a means of mitigating the pandemic. A wide variety of masks, ranging from cotton masks to N95 respirators, have been used by the general population throughout the pandemic. Intuitively, masks create a physical barrier between the wearer’s airway and the environment, reducing the chance of both disseminating and acquiring infection. In addition, a number of laboratory experiments, often based on manikin simulators, have demonstrated high efficacies of different masks including cotton masks; measured reductions of inhaled particles in laboratory settings ranged from ∼50% to >90% (1-3). However, the reported effectiveness of masking, demonstrated from randomized controlled trials (RCTs), has been much lower than the laboratory experiments, ranging from 10-25% (4, 5) to inconclusive in a 2023 Cochrane review (6). Further, many places have endured multiple COVID-19 pandemic waves since March 2020, despite mask mandates implemented in public settings. The large discrepancy between the *efficacy* (measured under ideal conditions of mask quality and correct use, e.g., in the laboratory) and *effectiveness* (measured under real-world, often imperfect conditions, e.g., in RCTs) of masking has prompted debates on the need for mask use in the general population (7, 8).

To explain this large discrepancy, studies have cited a number of factors. These mainly include compliance (e.g., the percentage of people actually wearing masks could be lower than the expected 100% among the test arm of an RCT), consistency (e.g., participants may not wear masks all the time, including the moment of exposure), and timing of implementation (e.g., there may be little circulation/presence of pathogen during an RCT to support demonstration of an effect)(8-10). However, it is difficult to test these factors in real-world settings and, as such, opinions and practices are divided – some dismiss the low effectiveness estimates from RCTs and continue to mask; others dismiss the high laboratory efficacies and refuse to mask. In addition, it remains unclear when and to what extent masking can be effective at the population level.

Here, to inform more effective individual preventive cares and public health measures, we develop an agent-based model that comprehensively accounts for individual masking behaviors and efficacies as well as the nonlinear transmission process, and test the combined impact of these processes at the population level. Using realistic inputs of mask efficacy (i.e., 50-80%, mean = 66%; Table S1) and contact data at the individual level, the model is able to reproduce the lower effectiveness (e.g., <25%) at the population level, as observed in real-world settings. Model results highlight the main factors contributing to the lower effectiveness, illustrate the complexity of measuring the underlying impact of masking due to nonlinear epidemic dynamics, and provide a more complete picture of the protective impact of masking (e.g., reducing repeated infections).

## RESULTS

### Overview of the agent-based model and analyses

The model used here has several key features. First, it explicitly simulates the transmission process to capture the contagious nature of infectious diseases. That is, individual risk of infection is determined by behavior (e.g., contact rate and mask preference) as well as the people with whom an individual interacts (e.g., the infection status of contacts). This structure depicts the connectivity and spread of infection across a population, which is likely a more realistic representation of the real world (vs. e.g., regression models). Second, the model separates household and non-household contact and transmission. This formulation not only allows modeling of the relative importance of household and non-household transmission based on contact data, but also allows modeling of specific masking behavior in each setting (e.g., masks are often not used within a household). Third, the model records the history of each infection and vaccination to account for immune protection and waning for each individual and, through the aforementioned inter-person interactions, the combined outcomes at the population level. Fourth, the model is age specific, which allows age-specific contact rates (e.g., higher among school-age children), efficacy of masking (e.g., higher for older adults), and disease severity (Table S1). This differentiation allows more realistic representation of population heterogeneity. Thus, unlike previous approaches, we more comprehensively capture individual-level effects, nonlinear epidemic dynamics, and overall impact at the population level.

Using the model, we simulated a COVID-19-like pandemic and subsequent epidemic waves in a population of 100,000 people, from March 1, 2020 (roughly the pandemic onset) to December 31, 2024 (roughly a 5-year period). The basic reproduction number was set to 2.6 – similar to estimates for the ancestral SARS-CoV-2 (11-14) that initiated the COVID-19 pandemic – in our main analysis using contact data from the POLYMOD study (15) and assuming an infection probability of 50% (10%) for physical (nonphysical) contact. To simulate pandemic dynamics, we used mobility data to adjust contact rates and vaccination data to account for vaccine protection, for all simulations. To test the impact of masking, we varied the masking rate (i.e., the fraction of population using masks, 10-100%) and the timing of masking (always wear masks outside the home vs. when prevalence is above 0.01% to 2%) and compared simulated epidemic outcomes to a baseline setting without masking.

### The model recapitulates masking effectiveness reported in RCTs

Fig 1 shows example simulated daily infection rates during the ∼5-year study period. Under the most vigilant scenario in which people choosing to wear masks (i.e., wearers) always do so outside the home, increasing the masking rate greatly reduces epidemic size; this reduction is consistent across all years (Fig 1A). However, even with a near 100% masking rate (i.e., all except for those aged <1 year use masks), epidemic waves, albeit of much smaller size, persist. In addition, the effectiveness decreases substantially, when masking is implemented only when population prevalence of infection is high (e.g., >1% in Fig 1B). It is further evident from Fig 2A, across a wide range of scenarios and time frames, there are many instances that estimated effectiveness (measured as the reduction in total number of infections compared to the baseline without masking) is <25% – the higher bound of estimates reported in RCTs – including instances with high masking rates (e.g., >80%) if masking is enacted late (see cells within the black polygons in Fig 2A).

**Fig 1.**
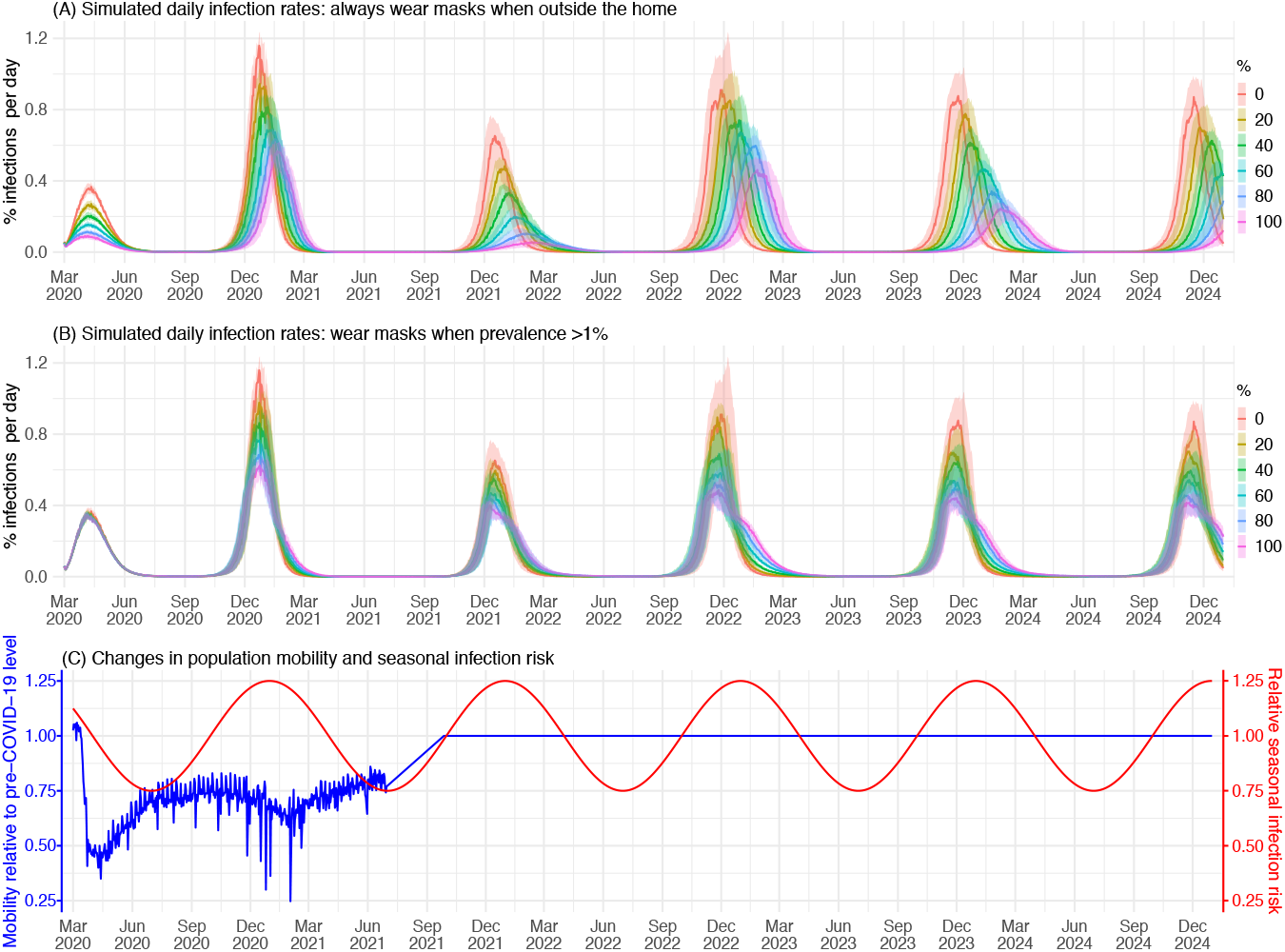
Example simulated epidemic infection rates. Colored lines (median) and shaded areas (interquartile ranges) show daily infection rates for simulated populations with different masking rates (see legend) and timing of masking: (A) mask wearers do so outside the home at all times or (B) only when infection prevalence is >1%. For context, (C) shows changes in daily population mobility (representing reductions in contact rate due to social distancing rules) and seasonal infection risk (representing effects due to environmental conditions), used in the simulations. Mobility data for New York City during March 2020 – June 2021 (shown here) were used directly as model inputs. From July 2021 onward, we assumed there was no social distancing and increased the mobility linearly to the pre-COVID level over a 3-month period and then remained at that level (i.e., set to 1). Trends in seasonal infection risk were represented using a sinusoidal function (see details in Methods).

**Fig 2.**
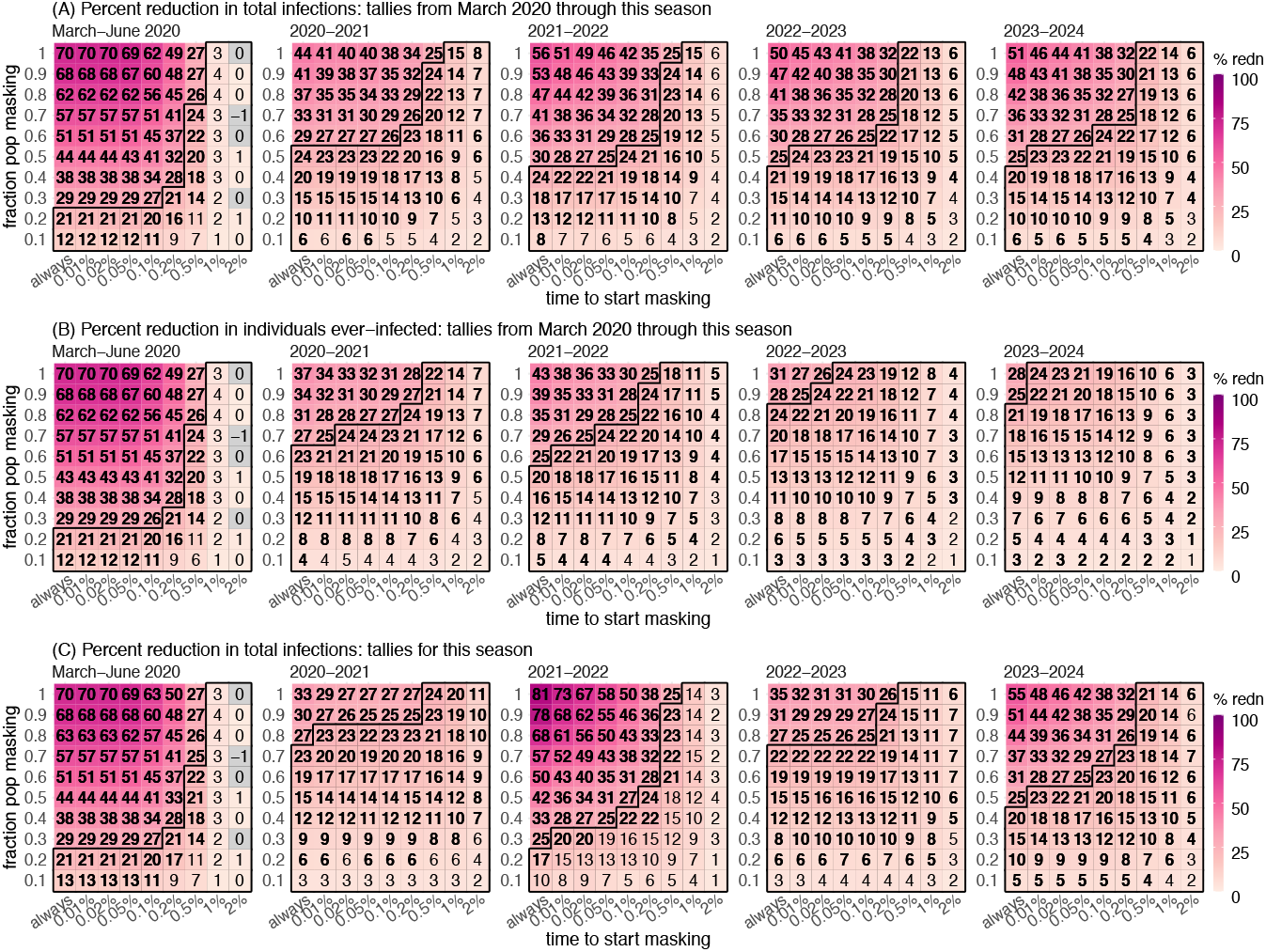
Estimated effectiveness of masking in reducing infection rates at the population level. Heatmaps show (A) percent reduction of cumulative total infections from March 2020 through different seasons (see panel subtitles, same below), (B) percent reduction of the cumulative fraction of population ever-infected, i.e., regardless of number of (re)infections, and (C) percent reduction of total infections tallied for each season. Here, a season runs one year from July 1 to June 30 of the following year. Numbers in the cells show median effectiveness estimates (computed per Eqn 2) for the corresponding masking rate (y-axis) and timing of masking (x-axis, percentages show population infection prevalence when masking is enacted). Bold fonts indicate the 2.5^th^% of the estimate is above 0 (i.e., significant at *a* = 0.05 level); note the 95% intervals here are likely narrower than measures in RCTs as the model assumes all infections are identified for the entire population. The black lines delineate cells with a median effectiveness estimate <25%.

### Factors contributing to lower masking effectiveness in the real world

To understand the main factors behind low masking effectiveness, we hypothesize that transmission may persist in household settings where contact tends to be greater, yet masking is rare. As shown in Fig 3A (contact rates formulated using the POLYMOD survey data (15)), for most individuals, household contact accounts for the majority of total contact, and are hence a likely principal source of infection. Thus, to test the impact of household transmission, we simulated a counterfactual setting in which people also wear masks at home (Fig 3B).

**Fig 3.**
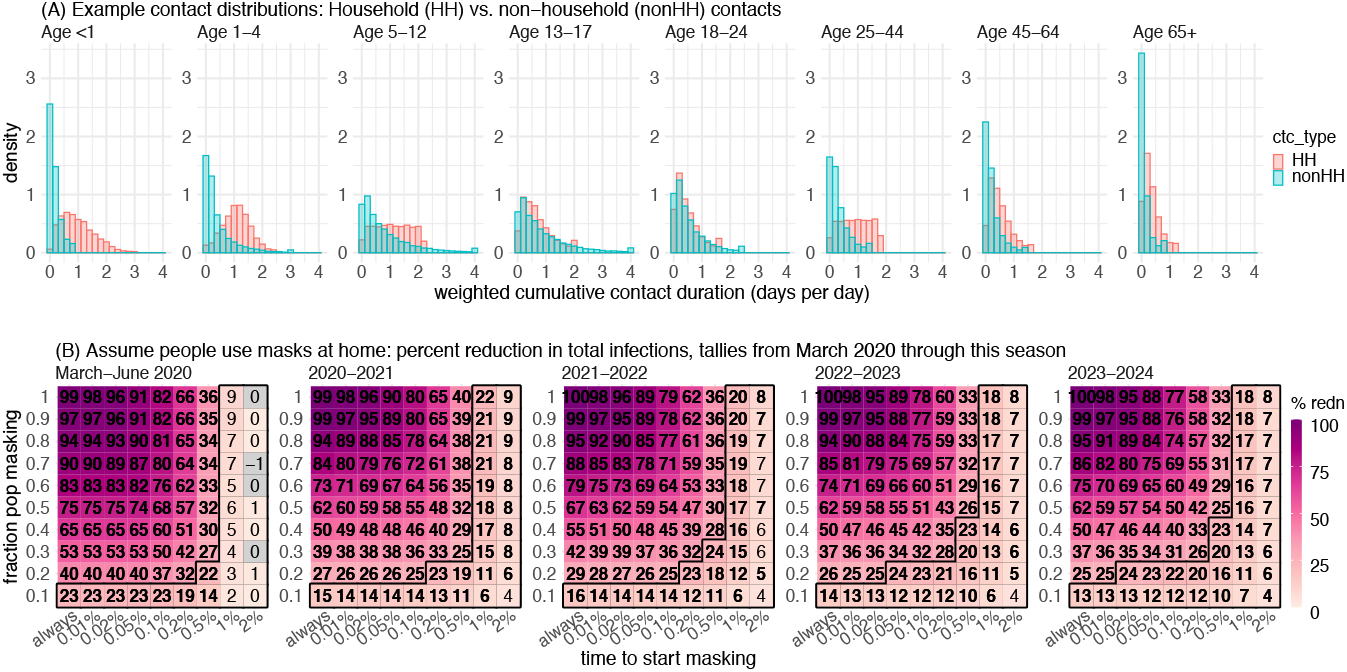
The impact of household transmission. (A) Example distributions of contact duration, formulated based on the POLYMOD survey (see details in Methods). (B) Counterfactual estimates of masking effectiveness in reducing total number of infections, assuming people universally wear masks at home. Numbers in the cells show the median effectiveness estimates (computed per Eqn 2) for the corresponding masking rate (y-axis) and timing of masking (x-axis, percentages show population infection prevalence when masking is enacted). Bold fonts indicate the 2.5^th^% of the estimate is above 0 (i.e., significant at *a* = 0.05 level); note the 95% intervals here are likely narrower than measures in RCTs as the model assumes all infections are identified for the entire population. The black lines delineate cells with a median effectiveness estimate <25%.

Three main findings emerge from the counterfactual analysis. First, compared to when masking is solely applied outside the home (main setting, Fig 2A), the effectiveness would be much greater should masking be also universally applied at home (counterfactual, Fig 3B). For instance, assuming a 50% masking rate and masking at all times, over the 5-year study period, the counterfactual effectiveness is 62%, more than twice of the estimate in the main setting (25%; Fig 2A, last panel, the cell in the 1^st^ column and middle row). Second, it is evident from Fig 3B, the effectiveness is higher than the expected direct effect should masking only affect the wearers. For instance, with a 50% masking rate and masking at all times, the expected effectiveness is 33% (i.e., 50% × 66% mean efficacy), much lower than the 62% estimate. This greater effectiveness highlights the indirect protection of masking to the entire population including non-wearers, an effect that is hidden in real-world settings. Third, there is often a delay in awareness of disease prevalence and in turn a delay in masking. Such delays further facilitate introduction of infection into households and onward transmission (both within and outside households), leading to decreases in the overall effectiveness over time. For instance, assuming a 50% masking rate and masking at all times, the estimated effectiveness in the main setting decreased from 44% during March – June 2020 to 25% at the end of the 5-year study period (Fig 2A, first vs. last panel), whereas the counterfactual estimates remained above 60% over the entire 5-year study period (Fig 3B, first vs. last panel).

The model results also illustrate how system nonlinear dynamics can affect masking effectiveness. For example, during the first wave (March – June 2020), estimated effectiveness surpasses the expected, under most combinations of masking rate and timing (Fig 2A, the number in the cell > masking rate × 66% mean efficacy). This higher-than-expected effectiveness comes from the transmission reduction due to masking, which, in combination with the reduction in contact rate and less conductive environmental conditions in late spring/summer (Fig 1C), reduces the effective reproduction number *R*_*e*_ to <1, preventing a large pandemic wave (note: *R*_*e*_ measures the average number of secondary infections and unity is the epidemic threshold). Conversely, when *R*_*e*_ is much higher than 1, the effectiveness tends to be lower than expected due to “leakage” via household transmission, as noted above. For instance, a large wave occurred during the 2020-2021 season (see Fig 1 A, 2^nd^ wave) under all masking rates, likely due to high population susceptibility and relaxed implementation of other NPIs (e.g., social distancing; Fig 1C, blue line) coinciding with more conducive transmission conditions during wintertime (Fig 1C, red line). As a result, the reduction in epidemic magnitude was much smaller than during the first wave. This is also evident in the cumulative effectiveness over time (Fig 2A, compare the higher effectiveness estimates for March-June 2020 in the 1^st^ panel to the lower effectiveness estimates for subsequent seasons in remaining panels).

### Measurement issues can further affect reported masking effectiveness

In addition to the factors noted above, the method and timing of measurement could both affect the reported effectiveness of masking. For the former, reported effectiveness would be lower than actual, if repeated infections are not counted. For instance, the number of people ever infected will inevitably increase over time. Studies that only measure the ever-infected while omitting reinfections will thus underestimate the actual (see the last 4 panels in Fig 2B that discount reinfections, vs. those in Fig 2A that include reinfections). For the latter, prior epidemic dynamics could affect outcomes measured during a given period and consequently obscure the true effectiveness for masking. As shown in Fig 1, epidemic size could vary across seasons, due to, e.g., accumulated population immunity. Studies that measure the effectiveness based on a specific time period without accounting for prior dynamics could thus obtain vastly different estimates (Fig 2C). These model results demonstrate potential measurement errors and highlight the importance of adopting more comprehensive measurements (e.g., including reinfections) and accounting for prior epidemic dynamics (e.g., examining cumulative effects).

### Additional factors (sensitivity analyses)

The main analysis above assumes that infection risk via nonphysical contact is 20% of that via physical contact (i.e., relative risk = 0.2). Depending on the etiologic agent, this relative risk could be higher or lower. In particular, higher infection risk via nonphysical contact may apply to pathogens that transmit more efficiently via the airborne route. Thus, we tested the model under different nonphysical/physical contact-mediated relative infection risks (0.1, 0.5, and 1, vs. 0.2 in the main analysis). Per the POLYMOD contact data, the basic reproduction number (*R*_*0*_, i.e., the average number of secondary infections in a naïve population) is slightly higher for a relative risk of 1 (3.2 vs. 2.5-2.6 for other settings). As expected, it is more challenging to use masking to reduce infection when nonphysical contact is as likely as physical contact to cause an infection (Fig S1, 1^st^ row same risk vs. other rows with lower relative risks). Nonetheless, the general findings noted in the main analysis are consistent with all other relative risk settings tested (Fig S1).

In addition, we hypothesize that mask wearers may be more risk adverse and thus may adopt additional precautionary measures such as reducing contact with others. We tested this by reducing the contact rate of mask wearers by 25%. As expected, the model estimates a further reduction in infections when mask wearers simultaneously reduce their contact rates (Fig S2 vs. Fig 2). For instance, the model estimates a 29% (vs. 23% in the main setting) reduction in total infections over 5 years, if half of the population use masks when prevalence is >0.01.

### Estimated effectiveness of masking at population and individual levels

To examine the effectiveness of masking at the population level, we compare simulated total numbers of infections in a population with different masking settings, to one with no masking. As shown in Fig 2A and Table S2, at the population level, the effectiveness of masking depends on both the masking rate and timeliness of implementation. When implemented early during the epidemic, masking can substantially reduce infections (e.g., ∼30% estimated reduction over the 5-year study period for a 70% masking rate), despite the “leakages” noted above. Model results using parameters based on COVID-19 also show similar reductions in hospitalizations (Fig S3 and Table S2), suggesting similar downstream impacts on severer disease outcomes. However, timely implementation is crucial and there appears to be a threshold, after which estimated effectiveness declines rapidly – e.g., when community prevalence is >0.2% for the COVID-19-like epidemics tested here (Table S2).

To examine the impacts of masking at the individual level, for each simulated population (under a specific masking setting), we compare the (re)infection rates among mask wearers with non-wearers. Across a wide range of settings tested, the model estimates substantial lower risk of reinfection for mask wearers (Fig 4A). For instance, the model estimates a ∼50% lower risk of having ≥3 infections for mask wearers, over the 5-year study period. In addition, stratifying by mask efficacy, the estimated protection against infection increases with mask efficacy, and this dose-response is evident across all age groups (Fig 4B) despite different contact patterns (Fig 3A).

**Fig 4.**
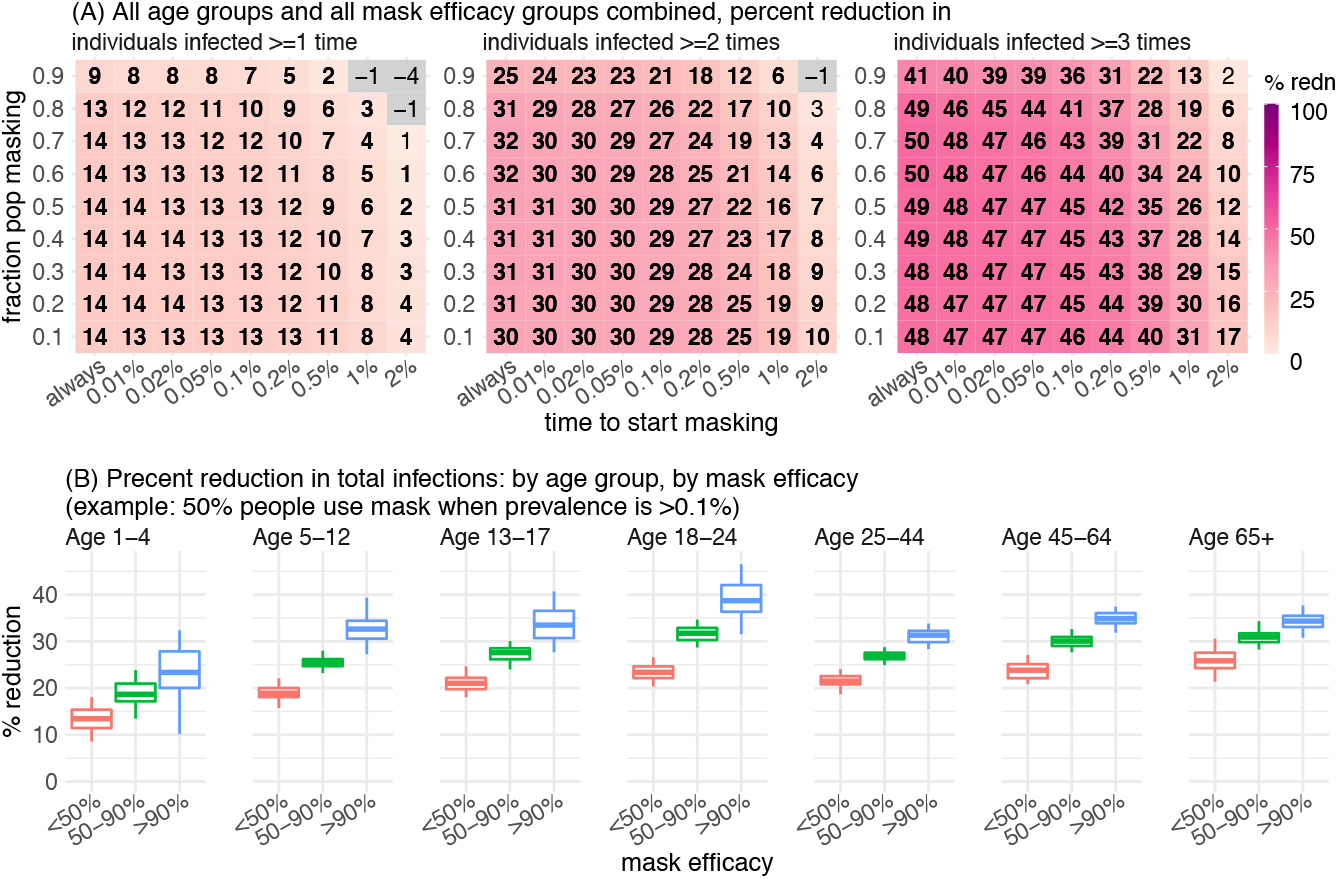
Estimated effectiveness of masking at the individual level. (A) Estimated percent reduction of repeated infections over the 5-year study period (see panel subtitles), comparing mask wearers to non-wearers in the same simulated population. Numbers in the cells show the median effectiveness estimates (computed per Eqn 3) for the corresponding masking rate (y-axis) and timing of masking (x-axis, percentages show population infection prevalence when masking is enacted). Bold fonts indicate the 2.5^th^% of the estimate is above 0 (i.e., significant at *a* = 0.05 level); note the 95% intervals here are likely narrower than measures in RCTs as the model assumes all infections are identified for the entire population. (B) Estimated percent reduction of total infections over the 5-year study period, by age group and mask efficacy. Boxplots show estimated median (middle bar), interquartile range (box edges), and 95% intervals (whiskers).

## DISCUSSION

Despite the high efficacy of masks in reducing particle inhalation, reported effectiveness of masking has been less than ideal. This large discrepancy has puzzled researchers and prompted doubt on the validity of masking as a public health measure. Here, our model not only generates consistent results reconciling the efficacy-effectiveness discrepancy, but also helps elucidate the underlying drivers. Model results identify factors that can lower effectiveness and highlight a key “leakage” source via household transmission. Further, we show the interaction of nonlinear infectious disease transmission dynamics, control measures (e.g., masking and contact reduction/social distancing), and resulting potential measurement biases. As such, the model findings here help provide a more complete picture of masking at both the individual and population levels.

For individuals, modeling results here clearly demonstrate that consistent masking can reduce the risk of initial infection, and, over time, reduce the frequency of repeated infection and hence the number of reinfections. For SARS-CoV-2/COVID-19, there remains large uncertainty in the health impact of reinfections. While population-level data have suggested a likely lower severity for reinfections due to prior immune protection (e.g., data from South Africa (16)), individual (re)infection severity may vary vastly. For some individuals, there may be a substantial risk of long-lasting post-acute sequelae following SARS-CoV-2 infection (i.e., long-COVID (17)) and the risk of long-COVID may further increase for subsequent reinfections. In light of the unknown long-term health impact of repeated infections, continued precautionary measures (e.g., masking) that reduce repeated infections may be advisable given these potential health impacts.

At the population level, model results here also demonstrate that masking can provide direct protection to mask wearers as well as indirect protection to non-wearers, collectively reducing epidemic intensity. In addition, as for other intervention measures, timely implementation of masking is crucial to ensure its effectiveness. However, consistent with the RCTs, model results here also indicate the overall effectiveness is likely often lower than the reported efficacy measured in laboratory settings. As demonstrated in our counterfactual modeling and testing of a wide range of masking settings, the ultimate effectiveness is limited by multiple factors (particularly, household transmission). It is thus important to recognize this less-than-ideal effectiveness, in order to more effectively devise public health measures (e.g., planning for additional preventive measures).

To focus on the general impact of masking and influencing factors, here we sidestepped several processes shaping SARS-CoV-2/COVID-19 dynamics; these include the emergence and circulation of different variants throughout the pandemic, differential protection of vaccines against different variants, and potential changes in disease severity for subsequent repeated infections. In particular, for more infectious variants (higher *R*_*0*_), it may be more challenging to reduce transmission via masking, as preliminarily shown in our sensitivity analysis (Fig S1). However, the model developed here provides a general framework to study these questions and beyond.

Despite these limitations, the quantitative modeling developed here helps tease apart the various factors affecting masking effectiveness in the real world. Overall, our results suggest it is important for individuals to use masks during an epidemic, and for policy makers to recognize the nonideal effectiveness of masking when devising public health measures.

## METHODS

### Data sources and processing

Here we used contact survey data from the POLYMOD study (15, 18) to parameterize individual contact patterns (Fig 2A), and mobility data (Fig 1C) and vaccination data (Fig S4) from New York City (NYC) to model social distancing and vaccination during the pandemic. The POLYMOD study recorded contact data for 7290 participants (97,904 contacts) in 8 European countries (e.g., for each contact, household vs. non-household, physical vs nonphysical, and duration)(15, 18). To compute the individual contact duration, we pooled data of all participants, weighted nonphysical contact (multiplicatively by a factor 0.2 in the main analysis and 0.1, 0.5, or 1 in the sensitivity analysis) to account for likely lower per-contact infection risk than physical contact. We then summed the weighted contact durations by contact type (household vs. non-household) for each individual. Note that, an individual’s total contact duration per day could exceed one day because simultaneous contacts are counted separately and additively combined. Given reported differences by age group, we further stratified by age group, and fit the age-grouped, type-specific contact durations to a probability distribution to formulate the agent-based model. Briefly, for each age group, we fitted the data to six distributions (i.e., uniform, exponential, normal, log-normal, Weibull, or gamma) and selected the best-fit distribution based on the Bayesian information criterion (19). The specific distributions used are shown in Table S3.

During the COVID-19 pandemic, many places implemented social distancing policies to reduce population contact rates. To represent these dynamics, as in our previous work (20-22), we used mobility data derived from Google Community Mobility Reports (23) to model contact reductions. Here, we used data for NYC during March 2020 – June 2021 to formulate the model. From July 2021 onward, we assumed there was no social distancing and increased the mobility linearly to the pre-COVID level over a 3-month period and remained at that level for the remainder of the study period (Fig 1C).

In addition, vaccines became available in late December 2020 and mass vaccination was rolled out subsequently. To represent the impact of vaccinations, we used data from NYC (4 doses, 12/14/2020 – 3/9/2023, at the time of this study; Fig S4)(24) to formulate the model. The 1st and 2^nd^ doses were administered 3-4 weeks apart as the primary series and the 3^rd^ and 4^th^ doses were administered more than 6-12 months later. As there is likely little overlap of immune protection from the preceding vaccination series/dose, we additively combined vaccinations of the primary series, 3^rd^, and 4^th^ doses to formulate the daily vaccination rate in the model. Assuming similar vaccination uptakes in the future, we used the mean for the same day of the year over 3/1/2022 – 3/9/2023 for those under 25 (roughly a 1-year period) and 9/1/2021 – 3/9/2023 for those ≥25 years (longer period given the earlier vaccination rollout for older ages) to formulate vaccination rates for days without data. For simplicity, we assumed a 90% vaccination effectiveness for all doses.

### The agent-based model

The agent-based model simulates individual infection and disease history, depending on age, contact pattern, individual preventive measures (e.g., masking behavior), prior infection and immune protection, vaccinations and immune protection, and immunity waning. In addition, at the population level, the model incorporates infection seasonality, mobility data to represent social distancing and reductions in contact during the pandemic, and vaccination data to represent mass vaccination during the study period. We describe each component below.

1. Risk of infection: Each day, infection risk for a susceptible individual is computed as the complement of the probability of avoiding infection by household and non-household contacts, i.e.,

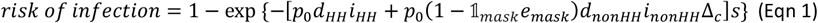

where *p*_*0*_ is the per-contact infection probability (set to 50% here); *d*_*HH*_ and *d*_*nonHH*_ are the total duration of household and non-household contact, respectively, formulated per the POLYMOD data, as described above. The infection prevalence within (*i*_*HH*_) and outside (*i*_*nonHH*_) household are computed by the model; here, for simplicity, we set *i*_*nonHH*_ to the population prevalence and *i*_*HH*_ to the mask-group-specific prevalence (i.e., assuming similar infection risk for households with similar masking behavior). For non-household contact, the term (1 − 𝟙_*mask*_*e*_*mask*_) represents the reduction in infection risk due to masking, with 𝟙_*mask*_ indicating whether the individual wears a mask outside (1 if yes and 0 otherwise) and *e*_*mask*_ being the mask efficacy (Table S1); Δ_*c*_ represents the further reduction through social distancing and was formulated using mobility data as described above (Fig 1C). The infection risk may further depend on one’s susceptibility (*s*), e.g., multiplicatively in Eqn 1; here, for simplicity, we set *s* to 1 for all ages.
2. Progression of infection: Infected (i.e., exposed but not yet infectious) individuals progress through the latency and infectious period exponentially, per: 1 − exp (−1/*D · d*), where *D* is the mean sojourn time (i.e., latency or infectious period; Table S1), and *d* the time since the corresponding transition.
3. Immune protection and waning: Due to limited data on immune protection, here we assumed immune protection wanes per a logistic function: 1/(1 + exp (−*k*(*τ* − 0.5*T*)), where *T* is the immunity period and *τ* the time since the corresponding event (i.e., infection or vaccination). To estimate the parameters (*k* and *T*), we fit the function to vaccination effectiveness duration data from the UKHSA (25) and scaled the estimates to represent the duration of protection against infection (here, *T* = 321 days for prior infection and 180 days for vaccination) and hospitalization (*T* = 321 days for prior infection and 385 days for vaccination). That is, we assumed shorter protection against infection but longer protection against severe disease from vaccination, and moderate protection against infection and severe disease from prior infection.
4. Vaccination: Using the daily vaccination rate computed above (Fig S4), we sampled individuals for vaccination according to their infection status (excluding those actively infectious) and vaccination probability based on their time since last vaccination and the estimated immunity period.
5. Hospitalization: For infected individuals, we computed the probability of hospitalization, based on the age-specific hospitalization rate (*h*_*a*_, Table S1) and immune protection from vaccination (*p*_*vac*.*immunity*_) and/or prior infection (*p*_*inf*.*immunity*_), per: *h*_*a*_(1 − 𝟙_*vaccinee*_ *· p*_*vac*.*immunity*_)(1 − 𝟙_*recoveree*_ *· p*_*inf*.*immunity*_). The level of immune protection *p*_*vac*.*immunity*_ and *p*_*inf*.*immunity*_ are computed using the aforementioned logistic function and corresponding immunity period estimates.
6. Seasonality: To represent the more conducive transmission conditions during winter months for respiratory viruses, including SARS-CoV-2 (26), we scaled the risk of infection (Eqn 1) by the seasonal risk (*b*_*s*_). For simplicity, here we modeled seasonality using a sinusoidal function: 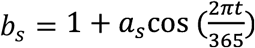, where *a*_*s*_ is the seasonality amplitude (set to 0.25 here) and *t* is the day of the year.
7. Demographics: The model simulates births and deaths (both with a rate set to 1/85 years to maintain the population size) by sampling and replacing those >65 years with newborns (age = 0), and simulates aging by increasing the age of all individuals by 1 at the end of each calendar year.

### Simulation settings and estimation of masking effectiveness

For all simulations, we initialized the model using 100,000 people and 100 initial infections on March 1, 2020, and ran the model for ∼5 years (from 3/1/2020 to 12/31/2024); we repeated each simulation 100 times to account for model stochasticity and computed summary statistics [median, interquartile range, and 95% interval (i.e., 2.5% and 97.5% percentile)]. For the main analysis (masking is applied only outside the home), to test the effectiveness of masking under different conditions, we varied the masking rate from 10% to 100% with a 10% increment and assigned masking choice for each individual (all those aged ≥1 year; i.e., no masking for those under 1) using random sampling. For each masking rate, we further varied the timing of masking, from masking outside the home at all times to only when prevalence is above 0.01%, 0.02%, 0.05%, 0.1%, 0.2%, 0.5%, 1% and 2%, separately. In total, there were 90 (10 masking rates × 9 timings) combinations. For comparison, we also simulated a baseline setting with no masking.

For the counterfactual analysis (masking is applied both within and outside the home), we computed the risk of infection as: 1 − exp [−*p*_0_ (1 − 𝟙_*mask*_*e*_*mask*_)(*d*_*HH*_*i*_*HH*_ + *d*_*nonHH*_*i*_*nonHH*_ Δ_*c*_)*S*], instead of using Eqn 1. For the 1^st^ sensitivity analysis, we used different weights to scale the relative per-contact infection risk for nonphysical contact (0.1, 0.5, or 1, vs. 0.2 in the main analysis). In the 2^nd^ sensitivity analysis, we tested a 25% additional reduction in contact rate for mask wearers, by setting Δ_*c*_ in Eqn 1 to 0.75Δ_*c*_ for individuals using masks.

At the population level, the effectiveness of masking is estimated per the reduction in a given health outcome (infection, repeated infection, hospitalization, or repeated hospitalization) under a given masking setting (*x*_*masking*_) compared to the baseline setting without masking (*x*_0_), i.e.,

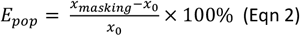

At the individual (group) level, we compare the rates of a given health outcome (*r*, i.e., normalized by the number of individuals in each group) among mask wearers versus non-wearers in the same simulated population, i.e.,

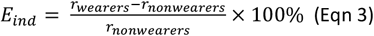

All inference, forecast, and statistical analyses were carried out using the R language (https://www.r-project.org).

## Data Availability

All data are publicly available as described in the Methods.

## Acknowledgements

This study was in part supported by the National Institute of Allergy and Infectious Diseases (AI145883 and AI163023) and the CDC Center for Forecasting and Outbreak Analytics (contract no.: 75D30122C14289).

## Supplemental Figures and Tables

**Fig S1.**
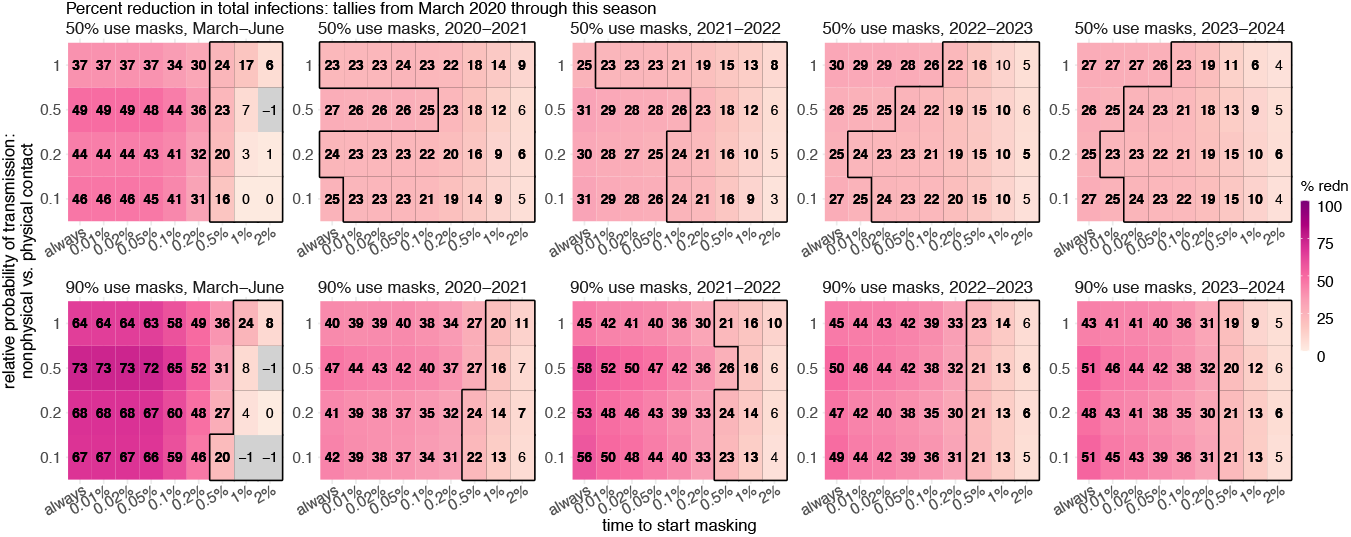
Estimated effectiveness of masking in reducing infection rates at the population level, assuming different relative infection risk by nonphysical contact compared to physical contact (y-axis). Panels in the top row assume 50% of the population use masks and the bottom row assume 90% of the population use masks. Other figure settings are the same as Fig 2A.

**Fig S2.**
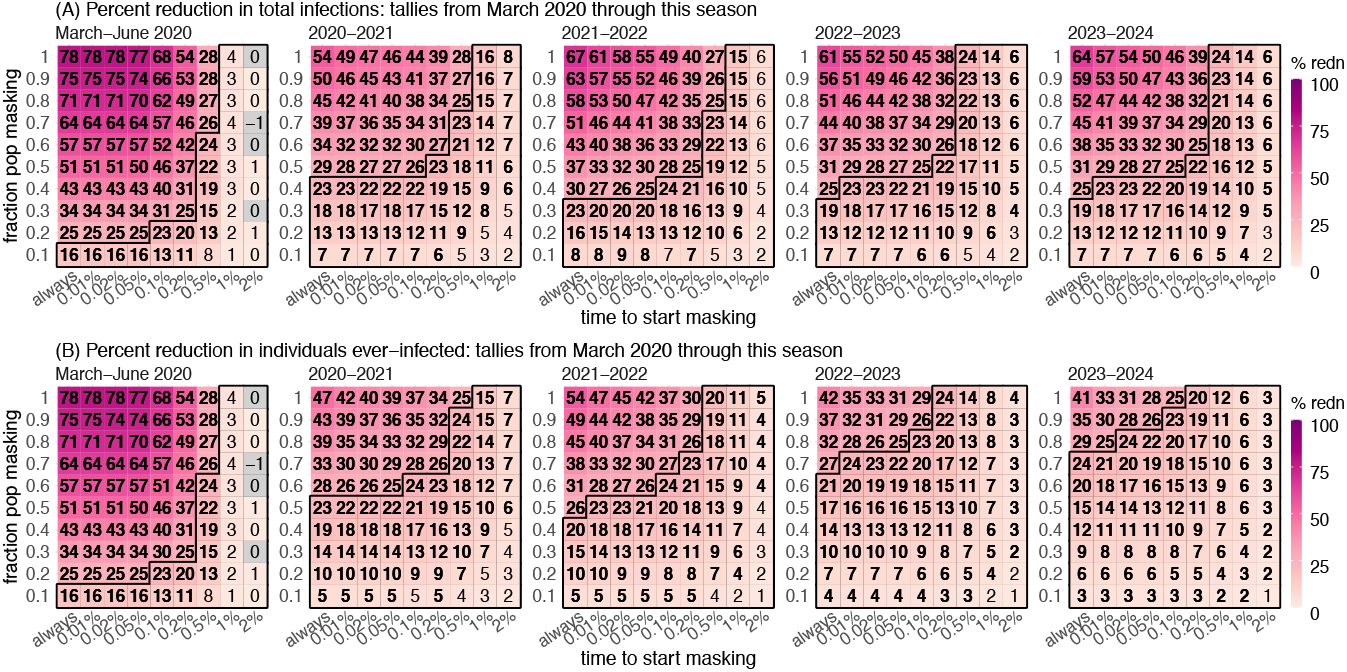
Impact of combinations with additional preventive measures. Model and figure settings are the same as Fig 2, except here the model assumes mask wearers additionally reduce their contact rate by 25%. Heatmaps show (A) percent reduction of cumulative total infections from March 2020 through different seasons (see panel subtitles) and (B) percent reduction of cumulative fraction of population ever-infected, both assuming mask wearers additionally reduce their contact rate by 25%. Here, a season spans July 1 to June 30 in the following year. Numbers in the cells show the median effectiveness estimates (computed per Eqn 2) for the corresponding masking rate (y-axis) and timing of masking (x-axis, percentages show population infection prevalence when masking is enacted). Bold fonts indicate the 2.5^th^% of the estimate is above 0 (i.e., significant at *a* = 0.05 level); note the 95% intervals here are likely narrower than measures in RCTs as the model assumes all infections are identified for the entire population. The black lines delineate cells with a median effectiveness estimate <25%.

**Fig S3.**
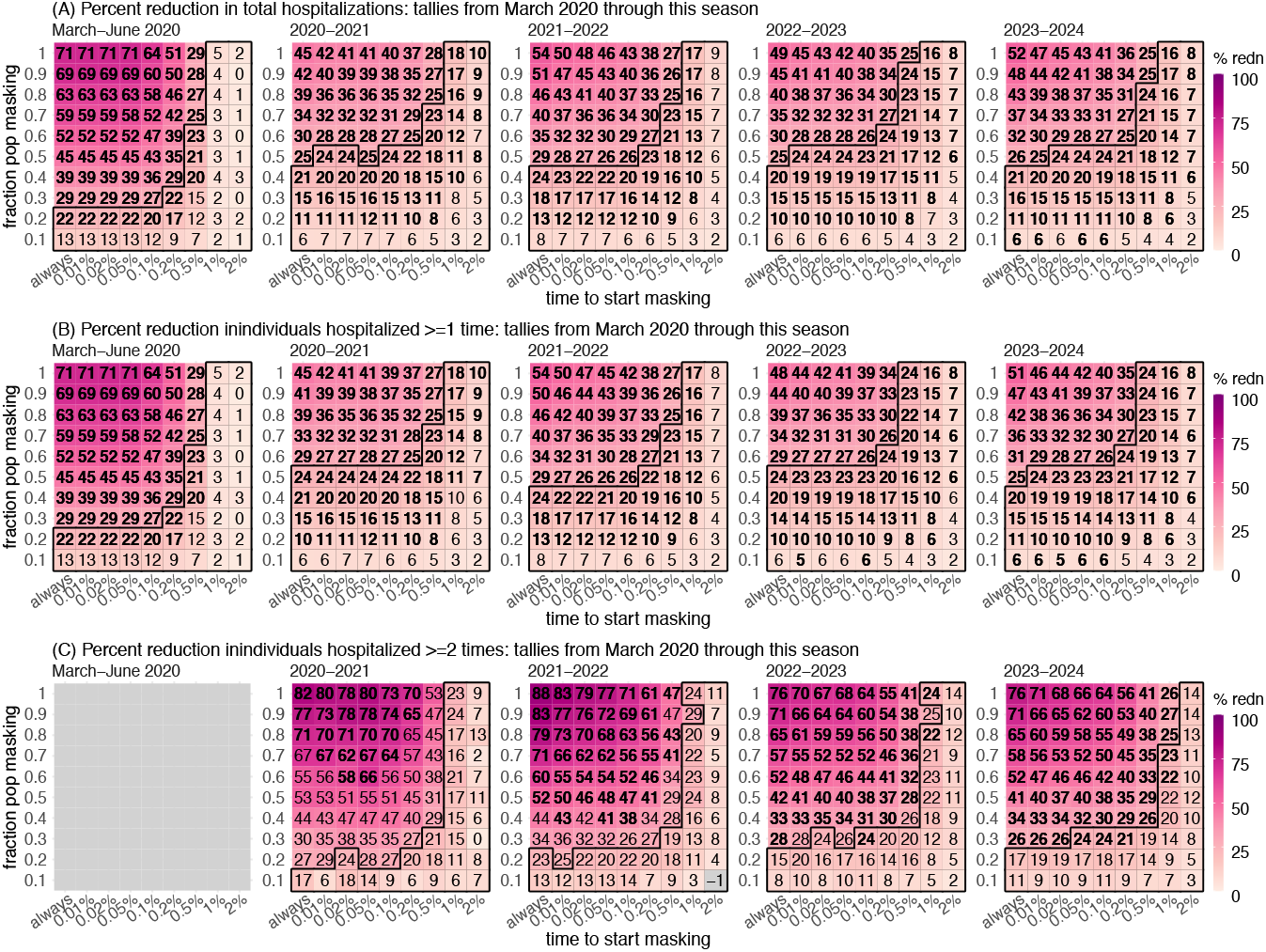
Estimated effectiveness of masking in reducing *hospitalizations* at the population level. Heatmaps show (A) percent reduction of cumulative total hospitalizations from March 2020 through different seasons (see panel subtitles, same below), (B) percent reduction of cumulative fraction of population hospitalized at least once, and (C) percent reduction of cumulative fraction of population hospitalized more than once (March-June 2020 is not shown given the lack of repeated hospitalizations during the short time period). Here, a season spans from July 1 to June 30 in the following year. Numbers in the cells show the median effectiveness estimates (computed per Eqn 2) for the corresponding masking rate (y-axis) and masking (x-axis, percentages show population infection prevalence when masking is enacted). Bold fonts indicate the 2.5^th^% of the estimate is above 0 (i.e., significant at *a* = 0.05 level); note the 95% intervals here are likely narrower than measures in RCTs as the model assumes all hospitalizations are identified for the entire population. The black lines delineate cells with a median effectiveness estimate <25%.

**Fig S4.**
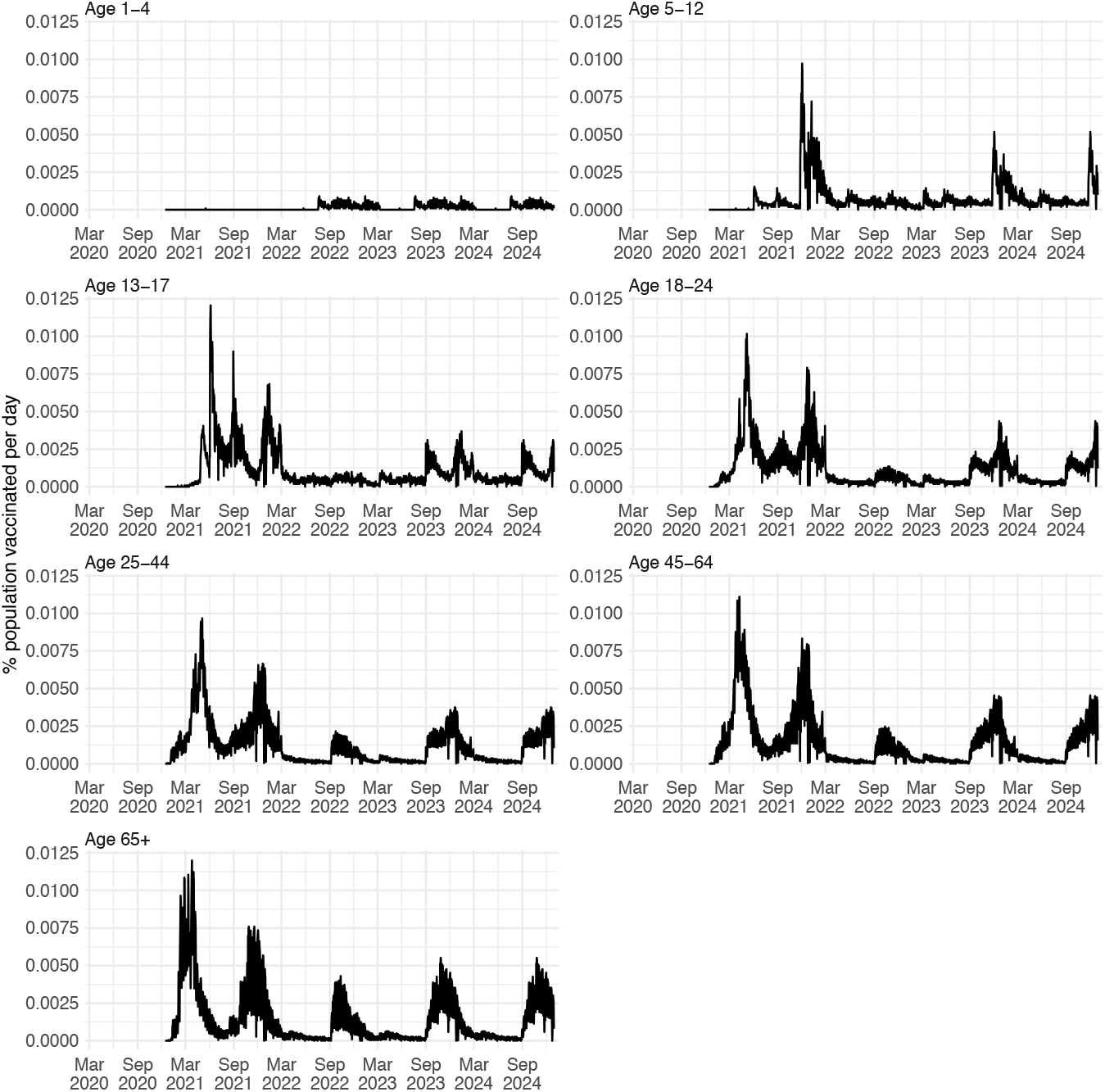
Daily vaccination rates used in the simulations. Data for New York City during 12/14/2020 – 3/9/2023 were used directly as model inputs. For days without data, we used the mean for the same day of the year over 3/1/2022 – 3/9/2023 for those under 25 (roughly a 1-year period) and 9/1/2021 – 3/9/2023 for those ≥25 years (longer period given the earlier vaccination rollout for older ages).

**Table S1.** Parameter values used in simulations.

| Parameter | Age<br>(year) | Value |
| --- | --- | --- |
| Initial number of infections | NA | 100 |
| Daily seeding rate | NA | 0.1 |
| Vaccination Effectiveness (VE) | NA | 90% |
| Latency period | NA | 3 days |
| Infectious period | NA | 5 days |
| Immune duration of prior infection against infection | NA | 321 days |
| Immune duration of prior infection against severe disease | NA | 321 days |
| Immune duration of vaccination against infection | NA | 180 days |
| Immune duration of vaccination against severe disease | NA | 385 days |
| Mask efficacy (age specific) | <1 | no masks |
| Mask efficacy (age specific) | 1-4 | 50% (SD = 25%) |
| Mask efficacy (age specific) | 5-12 | 50% (SD = 25%) |
| Mask efficacy (age specific) | 13-17 | 50% (SD = 25%) |
| Mask efficacy (age specific) | 18-24 | 50% (SD = 25%) |
| Mask efficacy (age specific) | 25-44 | 70% (SD = 25%) |
| Mask efficacy (age specific) | 45-64 | 70% (SD = 25%) |
| Mask efficacy (age specific) | 65+ | 80% (SD = 25%) |
| hospitalization rate (age specific) | <20 | $5 \times 10^{-4}$ |
| hospitalization rate (age specific) | 20+ | $\exp(-7.5 + 0.075\text{Age})$ |

**Table S2.** Model-estimated effectiveness of masking in reducing total infections and hospitalizations from March 2020 through the 2023-2024 season. Numbers (in percentage, %) show the estimated median (and 95% intervals) under different masking settings (see 2^nd^ column for the masking rate in the population and the columns 3-9 for the timing of masking). Note the 95% intervals here are likely narrower than measures in RCTs as the model assumes all infections and hospitalizations are identified for the entire population.

| Health outcomes | Fraction of population masking | Time to start masking |  |  |  |  |  |  |
| --- | --- | --- | --- | --- | --- | --- | --- | --- |
|  |  | Always | Prevalence >0.01% | Prevalence >0.02% | Prevalence >0.1% | Prevalence >0.2% | Prevalence >1% | Prevalence >2% |
| Infections | 0.1 | 5.9 (2.4, 8.7) | 5.4 (2.4, 8.6) | 6 (1.4, 9) | 5.3 (1.8, 9.3) | 5.2 (1.1, 9.5) | 3.4 (-0.5, 6) | 1.9 (-1.6, 5.9) |
| Infections | 0.2 | 10.5 (7.4, 13.7) | 9.8 (6.3, 14.2) | 9.7 (6.9, 14.2) | 9.3 (4.7, 12.7) | 8.6 (5.1, 12.7) | 5.4 (2.1, 8.8) | 2.9 (-0.7, 6.1) |
| Infections | 0.3 | 15.3 (10.2, 21.5) | 14.2 (11.4, 18.4) | 14.6 (10.3, 17.3) | 13.2 (9.4, 16.5) | 12.2 (8.4, 15.2) | 7.4 (3.9, 10.6) | 4.3 (0.4, 7.1) |
| Infections | 0.4 | 20.4 (16.3, 25.3) | 18.9 (15.5, 22.5) | 18.5 (14.9, 21.6) | 16.9 (13, 20.3) | 15.7 (12.2, 19.6) | 9.1 (5.2, 12) | 4.3 (0.8, 8.4) |
| Infections | 0.5 | 25.3 (21.6, 29.8) | 23.4 (19.9, 27.8) | 22.8 (19.2, 27) | 20.9 (17.2, 24.6) | 18.6 (15.4, 21.9) | 10.3 (7.1, 14.1) | 5.5 (1.8, 9.2) |
| Infections | 0.6 | 30.5 (25.8, 38) | 27.8 (24.4, 32.7) | 27.4 (23.5, 30.6) | 24.4 (20.9, 27.8) | 21.6 (18.8, 24.9) | 11.6 (7.1, 15.2) | 5.6 (1.9, 8.3) |
| Infections | 0.7 | 36.1 (31.4, 44) | 33 (28.9, 36.7) | 31.5 (28.7, 36.7) | 28 (25.3, 32.2) | 24.7 (21.3, 29.3) | 12.1 (7.7, 15.6) | 5.7 (2, 9.5) |
| Infections | 0.8 | 42.3 (37.8, 50.8) | 37.8 (34.7, 42) | 36.3 (32.8, 39) | 31.7 (28.2, 34.5) | 27.1 (23.6, 30.4) | 13 (8.1, 16.9) | 6.1 (1, 9.4) |
| Infections | 0.9 | 47.9 (42.7, 56.5) | 42.6 (38.3, 46.2) | 40.6 (37.4, 45.5) | 34.9 (32.5, 38.3) | 30 (26.8, 32.7) | 13.4 (8.7, 17.6) | 6.2 (2.1, 9) |
| Infections | 1 | 51.2 (46.4, 63) | 45.9 (42.2, 50.4) | 43.8 (40.9, 47.2) | 37.8 (35.1, 41.1) | 31.8 (28.5, 35.3) | 13.7 (9.2, 17.8) | 6.4 (1.2, 10.6) |
| Hospitalizations | 0.1 | 6.4 (0.9, 11.1) | 5.7 (0.5, 10.7) | 5.7 (0, 10.6) | 5.7 (0.3, 11) | 5.5 (-1.8, 10) | 3.6 (-2.4, 7.4) | 2.2 (-3.8, 8.3) |
| Hospitalizations | 0.2 | 11.3 (6.3, 15.5) | 10.4 (5.5, 16.2) | 10.8 (4.8, 16.6) | 10.8 (4.9, 14.6) | 9.5 (4.2, 14.2) | 6.3 (1.4, 10.6) | 2.9 (-1.9, 8.3) |
| Hospitalizations | 0.3 | 16.2 (9.9, 22.1) | 15.4 (9.3, 20) | 15.4 (10, 19.4) | 14.7 (9, 20) | 13.5 (7.6, 18.7) | 8.4 (2, 14) | 4.6 (-0.9, 9.9) |
| Hospitalizations | 0.4 | 20.6 (14.8, 26.9) | 19.8 (14.6, 24.2) | 19.9 (14.1, 24.5) | 18.6 (13.6, 23.4) | 17.9 (12.5, 23.4) | 10.9 (4.9, 15) | 5.9 (0.2, 9.8) |
| Hospitalizations | 0.5 | 25.8 (21.2, 31.3) | 24.9 (19.4, 30.3) | 24.2 (19.3, 29.4) | 23.7 (17.9, 27.4) | 21.1 (16.6, 25.3) | 12.2 (7.6, 17.3) | 6.5 (1, 10.8) |
| Hospitalizations | 0.6 | 32.1 (25.9, 39.1) | 29.6 (24.2, 34.2) | 28.8 (24.1, 32.8) | 26.7 (22.4, 30.5) | 24.5 (19.8, 28.2) | 13.7 (8.9, 18.2) | 6.9 (1.4, 12.1) |
| Hospitalizations | 0.7 | 37.4 (31.7, 45.1) | 34.1 (29.8, 38.5) | 32.9 (28.4, 38.9) | 30.8 (26, 36.1) | 27.5 (23.1, 32.1) | 14.6 (9.4, 20.1) | 6.9 (2.2, 12.7) |
| Hospitalizations | 0.8 | 43.3 (38.1, 51.7) | 39 (34.9, 43.7) | 37.6 (33.2, 41.5) | 34.8 (30.3, 38) | 30.8 (26.1, 34.9) | 15.5 (9.1, 21.1) | 7.4 (1.9, 12.3) |
| Hospitalizations | 0.9 | 47.9 (42.7, 58.1) | 43.8 (38.8, 48.2) | 42.4 (38, 46.6) | 38.5 (33.5, 42) | 34 (29.3, 38) | 16.9 (11.3, 21.3) | 7.5 (1.2, 12.2) |
| Hospitalizations | 1 | 52.3 (46.1, 66) | 47.2 (43.2, 52.2) | 45.2 (41.3, 49) | 40.8 (36.3, 44.4) | 36.4 (30.5, 39.7) | 16.4 (10.7, 21.4) | 7.8 (2.8, 13.8) |

**Table S3.**
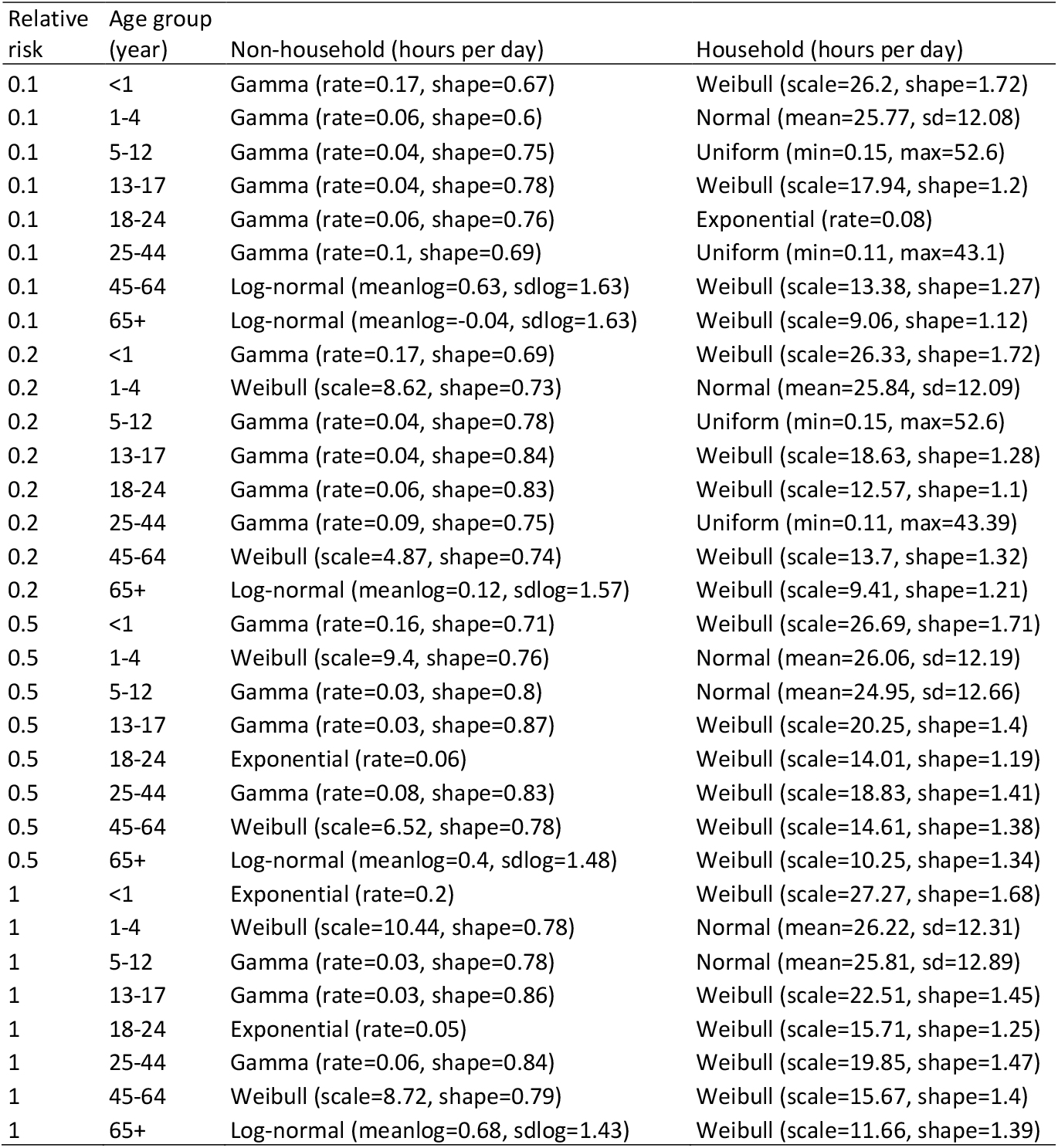
Probability distributions used to generate the individual contact durations in the agent-based model. We first weighted the POLYMOD contact duration data based on the relative infection risk by nonphysical versus physical contact; the weight was set to 1 for physical contact and 0.2 (main analysis), 0.1, 0.5, or 1 (sensitivity analyses; 1^st^ column) for non-physical contact. For each age group (2^nd^ column), we then fitted the weighted contact durations, stratified by contact type (non-household or household, 3^rd^ and 4^th^ columns, separately), to six distributions (i.e., uniform, exponential, normal, log-normal, Weibull, or gamma) and selected the best-fitting distribution based on the Bayesian information criterion.

| Relative risk | Age group (year) | Non-household (hours per day) | Household (hours per day) |
| --- | --- | --- | --- |
| 0.1 | <1 | Gamma (rate=0.17, shape=0.67) | Weibull (scale=26.2, shape=1.72) |
| 0.1 | 1-4 | Gamma (rate=0.06, shape=0.6) | Normal (mean=25.77, sd=12.08) |
| 0.1 | 5-12 | Gamma (rate=0.04, shape=0.75) | Uniform (min=0.15, max=52.6) |
| 0.1 | 13-17 | Gamma (rate=0.04, shape=0.78) | Weibull (scale=17.94, shape=1.2) |
| 0.1 | 18-24 | Gamma (rate=0.06, shape=0.76) | Exponential (rate=0.08) |
| 0.1 | 25-44 | Gamma (rate=0.1, shape=0.69) | Uniform (min=0.11, max=43.1) |
| 0.1 | 45-64 | Log-normal (meanlog=0.63, sdlog=1.63) | Weibull (scale=13.38, shape=1.27) |
| 0.1 | 65+ | Log-normal (meanlog=-0.04, sdlog=1.63) | Weibull (scale=9.06, shape=1.12) |
| 0.2 | <1 | Gamma (rate=0.17, shape=0.69) | Weibull (scale=26.33, shape=1.72) |
| 0.2 | 1-4 | Weibull (scale=8.62, shape=0.73) | Normal (mean=25.84, sd=12.09) |
| 0.2 | 5-12 | Gamma (rate=0.04, shape=0.78) | Uniform (min=0.15, max=52.6) |
| 0.2 | 13-17 | Gamma (rate=0.04, shape=0.84) | Weibull (scale=18.63, shape=1.28) |
| 0.2 | 18-24 | Gamma (rate=0.06, shape=0.83) | Weibull (scale=12.57, shape=1.1) |
| 0.2 | 25-44 | Gamma (rate=0.09, shape=0.75) | Uniform (min=0.11, max=43.39) |
| 0.2 | 45-64 | Weibull (scale=4.87, shape=0.74) | Weibull (scale=13.7, shape=1.32) |
| 0.2 | 65+ | Log-normal (meanlog=0.12, sdlog=1.57) | Weibull (scale=9.41, shape=1.21) |
| 0.5 | <1 | Gamma (rate=0.16, shape=0.71) | Weibull (scale=26.69, shape=1.71) |
| 0.5 | 1-4 | Weibull (scale=9.4, shape=0.76) | Normal (mean=26.06, sd=12.19) |
| 0.5 | 5-12 | Gamma (rate=0.03, shape=0.8) | Normal (mean=24.95, sd=12.66) |
| 0.5 | 13-17 | Gamma (rate=0.03, shape=0.87) | Weibull (scale=20.25, shape=1.4) |
| 0.5 | 18-24 | Exponential (rate=0.06) | Weibull (scale=14.01, shape=1.19) |
| 0.5 | 25-44 | Gamma (rate=0.08, shape=0.83) | Weibull (scale=18.83, shape=1.41) |
| 0.5 | 45-64 | Weibull (scale=6.52, shape=0.78) | Weibull (scale=14.61, shape=1.38) |
| 0.5 | 65+ | Log-normal (meanlog=0.4, sdlog=1.48) | Weibull (scale=10.25, shape=1.34) |
| 1 | <1 | Exponential (rate=0.2) | Weibull (scale=27.27, shape=1.68) |
| 1 | 1-4 | Weibull (scale=10.44, shape=0.78) | Normal (mean=26.22, sd=12.31) |
| 1 | 5-12 | Gamma (rate=0.03, shape=0.78) | Normal (mean=25.81, sd=12.89) |
| 1 | 13-17 | Gamma (rate=0.03, shape=0.86) | Weibull (scale=22.51, shape=1.45) |
| 1 | 18-24 | Exponential (rate=0.05) | Weibull (scale=15.71, shape=1.25) |
| 1 | 25-44 | Gamma (rate=0.06, shape=0.84) | Weibull (scale=19.85, shape=1.47) |
| 1 | 45-64 | Weibull (scale=8.72, shape=0.79) | Weibull (scale=15.67, shape=1.4) |
| 1 | 65+ | Log-normal (meanlog=0.68, sdlog=1.43) | Weibull (scale=11.66, shape=1.39) |

